# Impacts of bariatric surgery on costs and health outcomes of patients in Brazil: Interrupted time series analysis of multi-panel data

**DOI:** 10.1101/2020.08.14.20173708

**Authors:** José Antonio Orellana Turri, Edmund Chada Baraccat, José Maria Soarez, Lionai Lima dos Santos, Marco Aurélio Santo, Flavia Mori Sarti, Nana Kwame Anokye

## Abstract

**Study design:** Original research

Obesity is one of the most important public health problems worldwide, presenting significant socioeconomic impacts in health systems. Considering the rising costs of health care and the escalating burden of obesity in diverse countries, there has been increasing trends in examination of cost-effectiveness of health interventions towards prevention and treatment of obesity and its effects on comorbidities. Bariatric surgery has been considered an effective intervention for reducing moderate to severe obesity and improvement of obesity-related morbidities.

**Methodology:** Interrupted Time-Series Analysis (ITSA) on costs and health outcomes from retrospective cohort of 114 patients who had bariatric surgery at the Hospital of Clinics from the University of Sao Paulo. Medical records encompassing complete data on anthropometric, hemodynamic and biochemical parameters, utilization of resources and costs for health care procedures and regular assessments of patients’ health status associated with bariatric surgery at individual level were included in the study. Data on utilization of resources during outpatient and inpatient health care were used for estimation of patient’s direct costs referring to bariatric surgery, and 6-month pre- and post-intervention periods, adopting health system perspective in micro-costing approach.

**Results:** Mean direct costs of hospitalization (-US$2,762.22; −23.2%), image exams (-US$7.53; −0.8%) and medication (-US$175.37; −25,7%) presented decrease after bariatric surgery, and total direct cost (US$1,375.37; +138%), consultations (US$0.42; +2.4%) and laboratory exams (US$68.96; +63.4%) had increase. Reduction in weight, BMI, LDL, triglycerides, insulin, glucose-linked hemoglobin, and glucose showed improvements in patients’ health status after bariatric surgery. Cholesterol, VLDL, and HDL presenting increase after surgery.

**Conclusion:** Bariatric surgery represents an effective intervention for treatment of moderate to severe obesity with extensive benefits regarding health promotion and reduction of burden of disease. Trends in direct costs and multiple health outcomes showed post-intervention improvements in patients’ health status and reduction of health care needs of individuals.

## Introduction

Obesity is one of the most important public health problems worldwide nowadays, presenting significant socioeconomic impacts in national health systems. Higher prevalence of obesity has been associated with augmented onset of type 2 diabetes mellitus (T2DM), hypertension, and early mortality. Increases in Body Mass Index (BMI) that characterize overweight and obesity are associated with higher need for and expenditures with medications, primarily for the treatment of hypertension, diabetes and cardiovascular disease.^1–9^

Considering the rising costs of health care and the escalating burden of obesity in diverse countries, there has been increasing trends in examination of cost-effectiveness of health interventions towards prevention and treatment of obesity and its effects on comorbidities.^10–15^ Recent evidence on the impacts of obesity-related interventions, including prevention^16–18^ (promotion of physical activity and healthy eating) and treatment (medication and/or surgical procedures) strategies^19–24^, have demonstrated that efficacy of bariatric surgery with multi-disciplinary approach decreasing body weight and improving of health including cancer, cardiovascular events, T2DM, dyslipidemia, life expectancy, and quality of life.^5,23,25–28^ in the long run.^29–31^

Bariatric surgery has been considered an effective intervention for reducing moderate to severe obesity and improvement of obesity-related morbidities,^32^ being often used for its treatment in different countries, and has been included in the list of publicly funded procedures of some national health systems.^16–18^ Direct overall costs of bariatric surgery usually range from US$25,000 to US$30,000 per patient, and the annual health care costs for patients with BMI≥35kg/m^2^ generally vary between US$3,000 to US$10,000, in the treatment of T2DM, arterial hypertension and other.^3,5,33–35^

Considering that obese individuals present approximately doubled risk for utilization of medical services in comparison to eutrophic individuals (RR 1.89; CI95% 1.88-1.89, *p*<0.001), and mean annual medical costs are twofold higher in severe obesity (US$1,140 per person) relative to the general population (US$567 per person),^36^ the bariatric surgery can lead to cost savings estimated between US$2,016 to US$1,209 because of the decreased use of medication and reduction of events due to comorbidities.^27,37^

In Brazil, bariatric surgery was included in the list of procedures for treatment of moderate to severe obesity and made accessible to the general population within the Brazilian Unified Health System (Sistema Unico de Saude, SUS) in 1999.^38,39^ There has been increasing trends in adoption of bariatric surgery throughout the public facilities, especially among young females^38^, and results of previous studies have shown low mortality risk and high effectiveness of the surgery in the country.24,38,40–44

To date, most studies focusing on assessment of costs and effectiveness of bariatric surgery in Brazil and other countries have been based on limited sample size and/or single health outcome^5,6,23,38^, or relied on modelling of future outcomes. There is limited evidence including multiple anthropometric, hemodynamic and biochemical parameters of patients^20,22,45^, especially using longitudinal data before and after bariatric surgery^27,46^.

The absence of detailed information limits decision making around the use of bariatric surgery particularly given its high set up cost. In Brazil, there has been a significant increase in obesity rates in all age groups affecting in 2017 19% of the population. The Brazilian public health system offer freely bariatric surgery aiming to prevent possible complications related to obesity, but the lack of more accurate information about costs of the surgery, pre- and post-hospitalization and possible effectiveness can delay the spread of this procedure in the population.^47^ Data information on multiple health outcomes associated with health care costs of bariatric surgery throughout pre-surgery period until follow-up at individual level may provide important information for public policy decision making; particularly considering data from high complexity hospital considered reference health care institution in Brazil.^48–53^

The current study addresses the limitations in the literature by presenting an analysis of patient-level data in Brazil to examine the impact of bariatric surgery on multiple outcomes (including anthropometric, hemodynamic and biochemical parameters) using an interrupted time series approach. ITS is a useful quasi-experimental design for evaluation of longitudinal effects of interventions, especially based on natural experiments occurring in real-world settings.^54–60^

## Methodology

### Study design

Interrupted Time-Series Analysis (ITSA) on costs and health outcomes from retrospective cohort of patients who had bariatric surgery at the Hospital of Clinics from the University of Sao Paulo (HC-FMUSP), Brazil, from January to December of 2018.

ITSA regression model uses time series of particular outcome of interest to establish an underlying trend, which is interrupted by an intervention at a given known point in time. The statistical design of the model draws an expected trend in the hypothetical scenario without the intervention, comparing with the new trend established post-intervention in order to identify potential differences. The post-intervention scenario provides a comparison for evaluation of the intervention impact by calculating the change in slope throughout time, according to the following standard equation:^61,62^

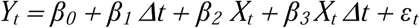

Where *Yt* is the accumulated result measured at each spaced time point *t*, *∆t* is the time since the start of the study, *Xt* is a dummy (indicator) variable representing the intervention (pre-intervention periods 0, or 1), and *Xt∆t* is an interaction term.^62^

### Bariatric surgery characteristics

The patients included in the study were distributed in three groups of surgery: Roux-en-Y gastric by-pass (R-YGB), vertical gastrectomy, and adjustable gastric banding. In most cases, open surgery was performed; however, a minor proportion of patients had surgery performed using video laparoscopy.

According to the standard procedures of bariatric surgery in Brazil, there is assessment of patients’ eligibility for bariatric surgery at primary health care facilities, and, depending on their health status, they are referred to a specialized health care unit. Patients with moderate to severe obesity diagnosis are referred to the Clinics Hospital, and included in the waiting list for bariatric surgery, performing pre-surgery monthly clinical and laboratory exams. After monitoring on eligibility criteria during variable period, patients are submitted to surgery, hospitalization and post-surgery follow-up, starting a post-surgical period of monthly clinic and laboratory exams (Figure 1).

**Figure 1.**
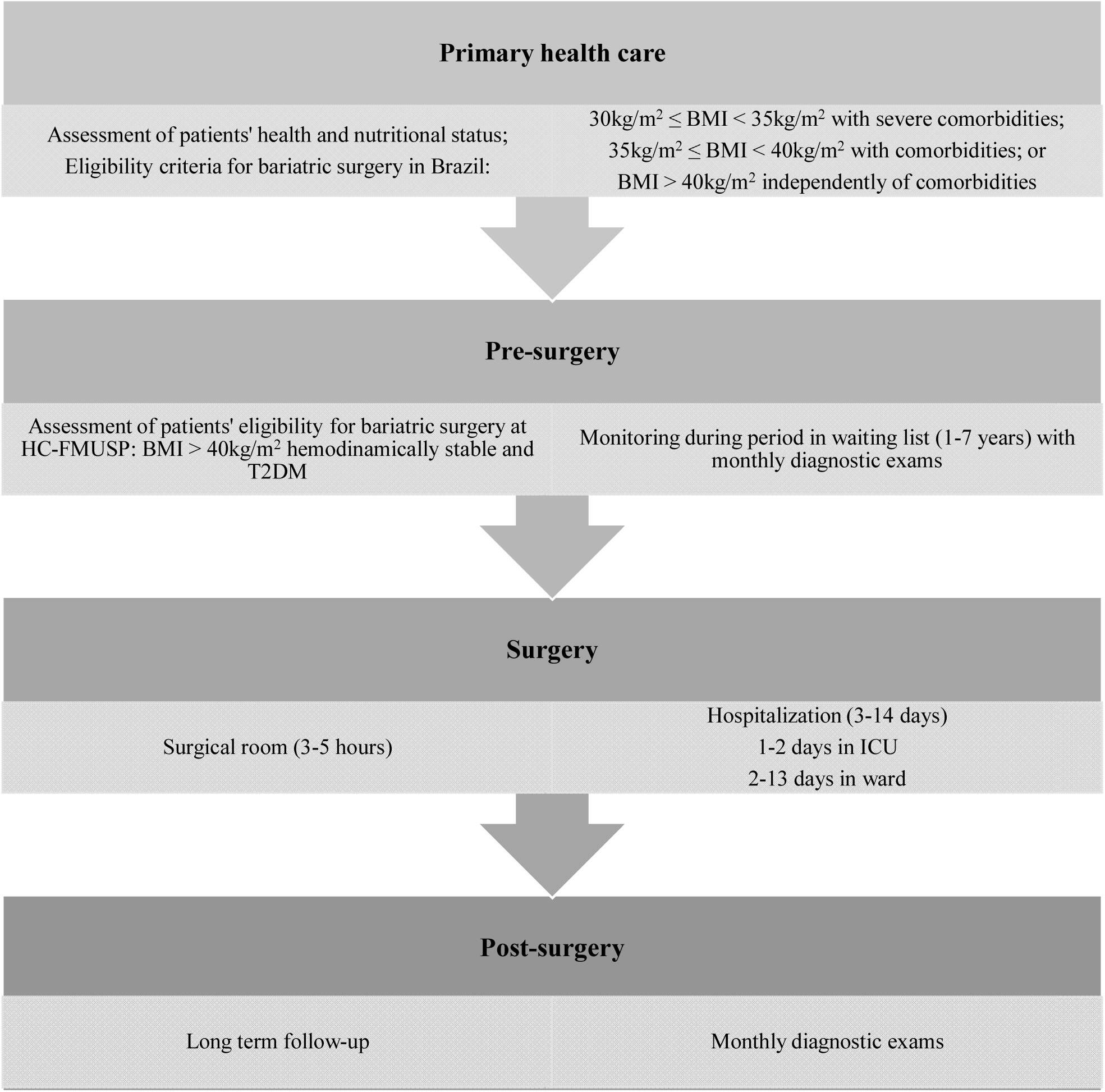
Flowchart of processes performed by patients eligible for bariatric surgery. Sao Paulo (Brazil), 2020. *Obs.: BMI = Body mass Index; T2DM = type 2 diabetes mellitus; ICU = Intensive Care Unit*.

### Sample

Information on cohort of 114 patients who had bariatric surgery at the HC-FMUSP in Sao Paulo, Brazil, through the Brazilian Unified Health System (SUS) from January to December of 2018, were obtained from hospital’s electronic medical records.

Only patients with medical records encompassing complete data on multiple anthropometric, hemodynamic and biochemical parameters, utilization of resources and costs for health care procedures and regular assessments of patients’ health status associated with bariatric surgery at individual level within the period of 6 months pre- and post-intervention (bariatric surgery) were included in the study.^63^ Demographic and lifestyle characteristics of patients were also gathered to comprise control variables in statistical analysis, including: gender and age; tobacco use and alcohol consumption.

Considering Brazilian protocols for bariatric surgery within SUS, patients are required to perform numerous exams and consultations pre- and post-surgery on regular basis.^39^ Individual information registered on daily-based electronic data collection at HC-FMUSP were gathered in single dataset referring to demographic and lifestyle characteristics, outpatient health care (pre- and post-intervention), and inpatient utilization of resources.

### Variables

Information of patients during the periods 180 days (6-month) pre- and post-intervention were obtained considering the baseline date (Table 1).

**Table 1.** Information on health outcomes and resources utilization of patients. Sao Paulo (Brazil), 2018

| Variable | Components |
| --- | --- |
| Health outcomes | Anthropometric measures (weight and height);<br>Hemodynamic measures (blood pressure);<br>Biochemical exams (cholesterol and fractions, triglycerides, insulin, glucose-linked hemoglobin, and fasting glucose). |
| Direct costs | Outpatient health care:<br>• Appointments with health professionals;<br>• Anthropometric measures (weight and height);<br>• Hemodynamic exams (blood pressure);<br>• Biochemical exams (cholesterol and fractions, triglycerides, insulin, glucose-linked hemoglobin, and fasting glucose).<br>Inpatient health care:<br>• Hospital length of stay;<br>• Type of surgery;<br>• Use of resources including: operating room, medication, meals, human resources, hemodynamic and biochemical exams. |

The criteria adopted for comparison of health outcomes and costs was measurement of changes in relation to the baseline of intervention (first day of hospitalization for bariatric surgery).

### Health outcomes

Information on health and nutritional status were obtained in electronic medical records of patients, referring to assessments pre- and post-surgery monitoring, considering its associations with bariatric surgery in the literature^5,23,25–31^; therefore, comprising the following set of health outcomes adopted in statistical analysis:

- Weight (kg);
- BMI (kg/m^2^);
- Blood pressure (mmHg);
- Cholesterol (mg/dL);
- VLDL (mg/dL);
- LDL (mg/dL);
- HDL (mg/dL);
- Tryglicerides (mg/dL);
- Insulin (IU/mL);
- Glucose-linked haemoglobin (%);
- Fasting glucose (mg/dL)

Measures were performed according to standard procedures internationally adopted within HC-FMUSP facilities by trained health professionals.

### Direct cost estimation

Data on utilization of resources during outpatient and inpatient health care were used for estimation of patient’s direct costs referring to bariatric surgery, and 6-month pre- and post-intervention periods, adopting health system perspective in micro-costing approach.

Prices of inputs and wages of health professionals involved in procedures were obtained within institutional database, based on information on recent purchases registered and human resources payroll. Prices per item were multiplied by the amount used for treatment of the patient, and hourly wages were multiplied by the amount of time dedicated to procedures and consultations performed during each patient’s treatment. Costs were updated to January 2020 and converted into US dollars using Brazilian Central Bank official exchange rate.

### Statistical analysis

Descriptive statistics and interrupted time series analysis (ITSA) with generalized estimating equations (GEE) and marginal effects were performed using single-center retrospective data on costs and multiple health outcomes related to bariatric surgery in patients from the HC-FMUSP. Information gathered for each patient was split into two segments for analysis, i.e., health outcomes and direct costs of health care before and after bariatric surgery, respectively.^63^

GEE with varying distribution were fitted for the different outcomes as follow: pre-intervention, intervention, post-intervention (gender and age) for adjustment of monthly trends according to patients’ characteristics. Marginal effects were obtained after GEE estimation by sample means at each period of evaluation (pre and post intervention) and are used for the incremental cost and effects over each outcome evaluated.

Dependent variables included in the models were direct costs of health care and health outcomes (weight, BMI, blood pressure, cholesterol and fractions, triglycerides, insulin, glucose-linked hemoglobin, and fasting glucose).

Interrupted time series ordered logistic were estimated for health outcomes, and Poisson models were estimated for health care costs, controlling for age and gender with random effects estimator. The interrupted time series model was achieved by defining independent variables *∆t* = time point (1,2,3,4,5,6,7,8,9,10,11,12,13), *X_t_* =0 for time points in stage 1 (time points 1, 2, 3, 4, 5 and 6) and 1 for time points in stage 2 (time points 7, 8, 9, 10, 11, 12 and 13), and *(XT)_t_* =0 for time points in stage 1, and (*t*−4) for time points in stage 2.

The regression coefficient for *T_t_* represents the rate of change of activity in stage 1, and the sum of regression coefficients for *T_t_* and *(XT)_t_* is the rate of change of activity in stage 2. Tthe effect over time was defined as the difference in the rate of change from stage 1 to stage 2, that is, the regression coefficient of *(XT)_t_*.

The immediate effect of the intervention was defined as the regression coefficient corresponding to *X_t_* (corresponding to the counterfactual difference between stage 1 and stage 2 evaluated at time point 7). The interrupted series time model for all costs and obesity-related measures was an ordinal logistic repeated measures model, including additive effects for *T_t_, X_t_* and *(XT)_t_*, and for all covariates to be adjusted for.

Statistical analysis were conducted using Stata version 14, and Newey-West standard errors were reported to account for autocorrelation at lag 1.^59^

### Patient and Public Involvement

Patients or the public were not involved in the design, or conduct, or reporting, or dissemination plans of this research.

### Ethical aspects

The study was approved by the Ethical Committee of the School of Medicine and Faculty of Public Health from the University of Sao Paulo (opinion numbers 0844918.6.3001.0068 and 90844918.6.0000.5421).

## Results

### Characteristics pre- and post-surgery

Data included information from 114 patients who had bariatric surgery in HC-FMUSP during 2018. Patients were previously enrolled in the waiting list for bariatric surgery at the hospital, and therefore were followed at least 180 days before and 180 days after the bariatric surgery. Considering characteristics at baseline, patients were approximately 48 years-old and the majority were female. Small proportion of individuals were smokers and most patients had comorbidities, especially hypertension and/or diabetes. The type of surgery conducted in most cases was open Y-Roux gastric by-pass (Table 2).

**Table 2.** Baseline characteristics of bariatric surgery patients. Sao Paulo (Brazil), 2018

| Gender |  | N | (%) |
| --- | --- | --- | --- |
| Men |  | 9 | 7.9% |
| Women |  | 95 | 83.3% |
| Age ($\mu \pm SD$) | | 47.8 ± 14.1 | |
| Lifestyle characteristics |  |  |  |
| Tobacco use |  | 20 | 17.5% |
| Alcohol consumption |  | 0 | 0.0% |
| Comorbidity |  |  |  |
| Hypertension |  | 72 | 63.2% |
| T2DM |  | 67 | 58.8% |
| Sleep apnea |  | 31 | 27.2% |
| Dyslipidemia |  | 25 | 21.9% |
| Arthrosis |  | 25 | 21.9% |
| Hypothyroidism |  | 10 | 8.8% |
| Varices |  | 10 | 8.8% |
| Chronic renal insufficiency |  | 10 | 8.8% |
| Cardiac Congestive Insufficiency |  | 5 | 4.4% |
| Type of surgery |  |  |  |
| Y-Roux gastric by-pass | Open | 67 | 58.8% |
|  | Video laparoscopic | 34 | 29.8% |
| Sleeve (vertical gastrectomy) | Open | 4 | 3.5% |
|  | Video laparoscopic | 3 | 2.6% |
| Adjustable gastric banding | Open | 3 | 2.6% |
|  | Video laparoscopic | 3 | 2.6% |

**Obs.:** $\mu$ = mean; SD = standard deviation; T2DM = type 2 diabetes mellitus

Mean direct costs of hospitalization (-US$2,762.22; -23.2%), image exams (-US$7.53; -0.8%) and medication (-US$175.37; −25,7%) presented decrease after bariatric surgery, and total direct cost (US$1,375.37; +138%), consultations (US$0.42; +2.4%) and laboratory exams (US$68.96; +63.4%) had increase in the same period, especially due to need of patients’ follow-up after intervention.

Regarding health outcomes, there were few obesity-related exams presenting increase after surgery (cholesterol, VLDL, and HDL); however, increase in HDL represents an improvement in health status. Similarly, reduction in weight, BMI, LDL, triglycerides, insulin, glucose-linked hemoglobin, and glucose showed improvements in patients’ health status after bariatric surgery (Table 3).

**Table 3.** Health outcomes and direct costs of patients during 6-month pre- and post-bariatric surgery. Sao Paulo (Brazil), 2018

| Health outcomes | 6-month period | | Difference | | Difference (US\$) | Cost per unit change (US\$) |
| --- | --- | --- | --- | --- | --- | --- |
| | Pre-surgery $\mu$ (±SD) | Post-surgery $\mu$ (±SD) | Mean | % | | |
| Weight (kg) | 125.53 (±18.58) | 103.23 (±24.46) | -22.3 | -17.8% | 22.3 | 61.68 |
| BMI (kg/m<sup>2</sup>) | 47.36 (±4.22) | 39.01 (±7.26) | -8.35 | -17.6% | 8.35 | 164.71 |
| Blood pressure (mmHg) | 132 (±15.21) | 126.51 (±17.36) | -5.49 | -4.2% | 5.49 | 250.52 |
| Cholesterol (mg/dL) | 165.11 (±39.9) | 167.78 (±38.55) | 2.67 | 1.6% | 2.67 | 515.12 |
| VLDL (mg/dL) | 21.51 (±8.33) | 22.91 (±8.93) | 1.4 | 6.5% | 1.4 | 982.41 |
| LDL (mg/dL) | 94.06 (±34.66) | 93.2 (±34.58) | -0.86 | -0.9% | 0.86 | 1599.27 |
| HDL (mg/dL) | 45.63 (±14.38) | 51.88 (±13.23) | 6.25 | 13.7% | 6.25 | 220.06 |
| Tryglicerides (mg/dL) | 130.67 (±64.21) | 117.27 (±56.95) | -13.4 | -10.3% | 13.4 | 102.64 |
| Insulin (IU/mL) | 17.19 (±22.55) | 12.69 (±7.47) | -4.5 | -26.2% | 4.5 | 305.64 |
| Glucose-linked haemoglobin (%) | 127.57 (±43.88) | 113.61 (±36.47) | -13.96 | -10.9% | 13.96 | 98.52 |
| Fasting glucose (mg/dL) | 113.22 (±45.77) | 99.11 (±39.83) | -14.11 | -12.5% | 14.11 | 97.47 |

| Health care costs | 6-month period | | Difference | | Difference US\$ | Cost per unit change |
| --- | --- | --- | --- | --- | --- | --- |
| | Pre-surgery $\mu$ (±SD) | Post-surgery $\mu$ (±SD) | Mean | % | | |
| Consultations | 17.64 (±13.9) | 18.06 (±15.32) | 0.42 | 2.4% | 0.42 | 3274.69 |
| Inpatient days | 11913.26 (±12682.01) | 9151.04 (±9695.16) | -2762.22 | -23.2% | 2762.22 | 0.50 |
| Laboratory exams | 108.72 (±112.94) | 177.68 (±217.78) | 68.96 | 63.4% | 68.96 | 19.94 |
| Image exams | 888.99 (±901.22) | 881.46 (±854.78) | -7.53 | -0.8% | 7.53 | 182.65 |
| Medication | 683.9 (±854.73) | 508.45 (±696.99) | -175.45 | -25.7% | 175.45 | 7.84 |
| Total direct costs | 996.61 (±4515.39) | 2371.98 (±6172.79) | 1375.37 | 138.0% | 1375.37 | - |

**Obs.:** $\mu$ = mean; SD = standard deviation. \*Including the cost of bariatric surgery

### Changes pre- and post-intervention using ITSA

Referring to results of the interrupted time series analysis, there was rise in overall costs at the period of intervention due to high costs of the bariatric surgery; however, it was followed by statistically significant decrease post-intervention. In general, post-surgery changes of health outcomes were also statistically significant, except cholesterol and LDL, showing improvements of patients’ health status after intervention (Table 4).

**Table 4.**
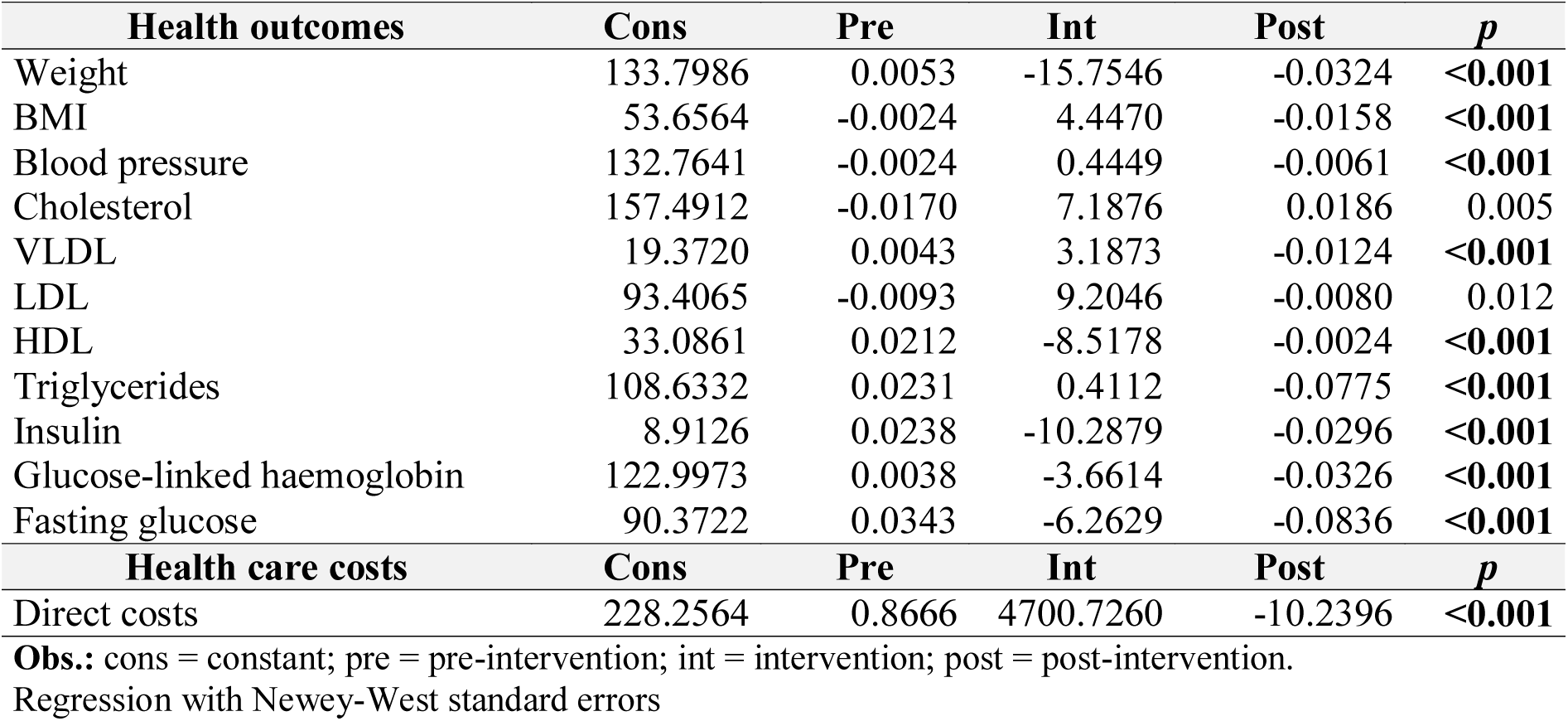
Pre- and post-intervention scenario of bariatric surgery using ITSA. Sao Paulo (Brazil), 2018

| Health outcomes | Cons | Pre | Int | Post | p |
| --- | --- | --- | --- | --- | --- |
| Weight | 133.7986 | 0.0053 | -15.7546 | -0.0324 | <0.001 |
| BMI | 53.6564 | -0.0024 | 4.4470 | -0.0158 | <0.001 |
| Blood pressure | 132.7641 | -0.0024 | 0.4449 | -0.0061 | <0.001 |
| Cholesterol | 157.4912 | -0.0170 | 7.1876 | 0.0186 | 0.005 |
| VLDL | 19.3720 | 0.0043 | 3.1873 | -0.0124 | <0.001 |
| LDL | 93.4065 | -0.0093 | 9.2046 | -0.0080 | 0.012 |
| HDL | 33.0861 | 0.0212 | -8.5178 | -0.0024 | <0.001 |
| Triglycerides | 108.6332 | 0.0231 | 0.4112 | -0.0775 | <0.001 |
| Insulin | 8.9126 | 0.0238 | -10.2879 | -0.0296 | <0.001 |
| Glucose-linked haemoglobin | 122.9973 | 0.0038 | -3.6614 | -0.0326 | <0.001 |
| Fasting glucose | 90.3722 | 0.0343 | -6.2629 | -0.0836 | <0.001 |
| Health care costs | Cons | Pre | Int | Post | p |
| Direct costs | 228.2564 | 0.8666 | 4700.7260 | -10.2396 | <0.001 |

**Obs.:** cons = constant; pre = pre-intervention; int = intervention; post = post-intervention.
Regression with Newey-West standard errors

### Post-intervention linear trends

Trends in post-intervention health outcomes showed reduction in most parameters, except cholesterol and HDL. In the case of HDL, increase in concentration indicates improvement in health status; whereas in the case of cholesterol, positive trend post-intervention was not statistically significant. BMI, VLDL, HDL, and fasting glucose showed important changes in trends pre- and post-surgery. Regarding direct costs, there was not a clear trend considering the 180-day period pre- and post-intervention (Table 5 and Figure 1).

**Table 5.** Post-intervention trends of health outcomes of bariatric surgery. Sao Paulo (Brazil), 2018

| Health outcomes | Trend | CI 95% | p |
| --- | --- | --- | --- |
| Weight | -6.2992 | (-0.0356;-0.0187) | <0.001 |
| BMI | -9.3199 | (-0.022;-0.0143) | <0.001 |
| Blood pressure | -3.0299 | (-0.0141;-0.003) | 0.0025 |
| Cholesterol | 0.2094 | (-0.0135;0.0168) | 0.8341 |
| VLDL | -4.9564 | (-0.0114;-0.0049) | <0.001 |
| LDL | -2.4414 | (-0.0312;-0.0034) | 0.0147 |
| HDL | 7.7052 | (0.014;0.0236) | <0.001 |
| Triglycerides | -4.0045 | (-0.0812;-0.0278) | <0.001 |
| Insulin | -3.5947 | (-0.009;-0.0027) | <0.001 |
| Glucose-linked hemoglobin | -3.4181 | (-0.0453;-0.0123) | 0.001 |
| Fasting glucose | -5.0545 | (-0.0683;-0.0301) | <0.001 |

**Obs.:** CI 95% = 95% confidence interval.
Regression with Newey-West standard errors.

**Figure 1.**
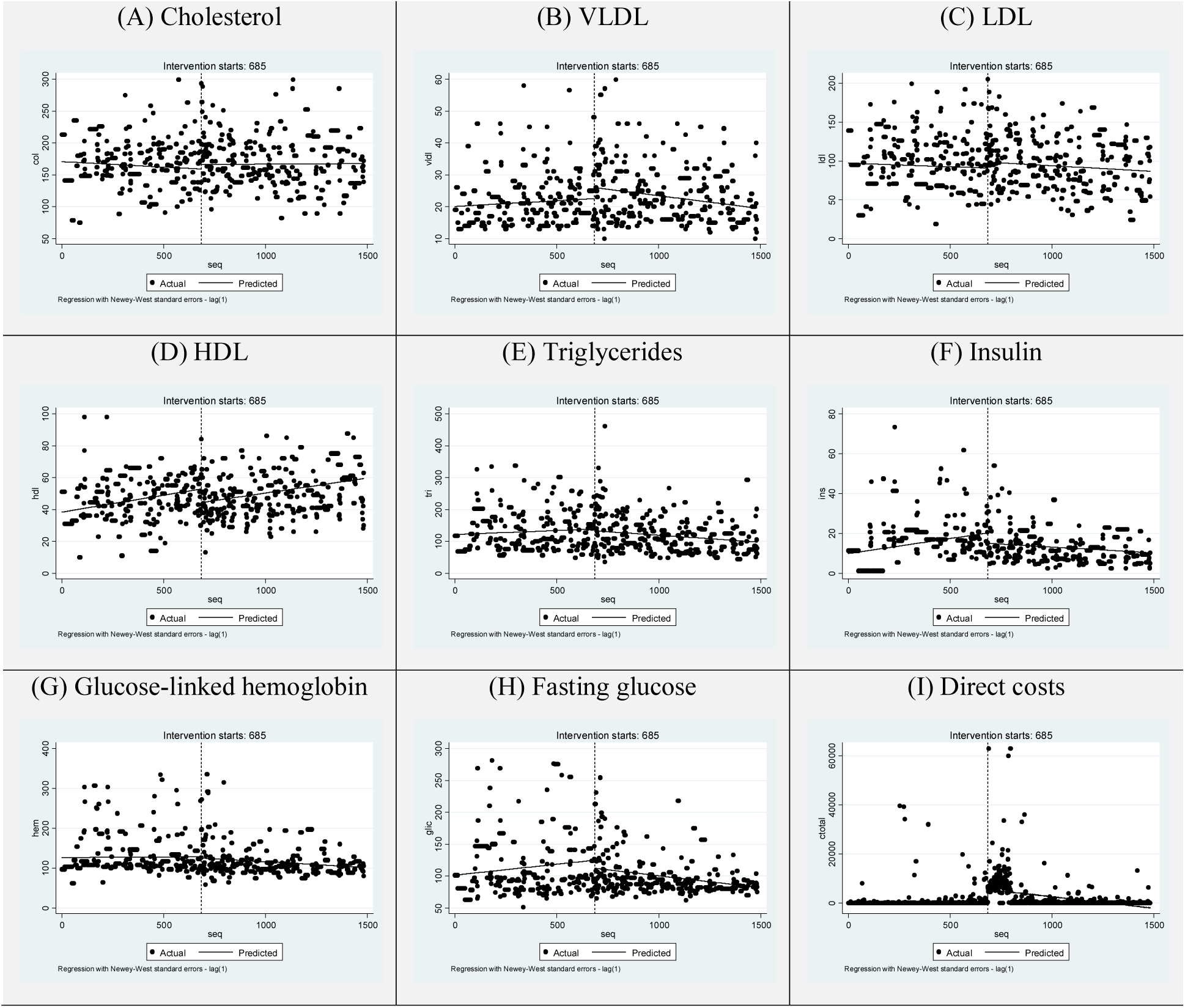
Trends of direct costs and health outcomes (laboratory exams) of bariatric surgery using ITSA for 180-day period pre- and post-intervention. Sao Paulo (Brazil), 2018

In addition, results of the generalized estimating multivariate regression showed that most changes observed in the comparison between pre- and post-surgery remain statistically significant controlling for patients’ characteristics. Marginal effects in direct health care costs post-intervention including covariates were significantly negative, similarly to health outcomes referring to blood pressure, VLDL, triglycerides, insulin, glucose-linked hemoglobin, and fasting glucose (Table 6).

**Table 6.** Marginal effects during pre-intervention, intervention, post-intervention adjusted by gender and age using GEE. Sao Paulo (Brazil), 2018

| Health Outcomes | Characteristics | dy/dx | CI95% | p |
| --- | --- | --- | --- | --- |
| BMI | Pre | 0.003 | (0.001 – 0.005) | <0.001 |
|  | Int | 0.803 | (0.006-0.154) | 0.033 |
|  | Post | -0.0006 | (-0.0008—0.0004) | <0.001 |
|  | Gender (female) | -0.005 | (-0.034—0.024) | 0.744 |
|  | Age | 0.0007 | (2.855-3.001) | 0.105 |
| Weight | Pre | 0.002 | (-0.01-0.01) | 0.804 |
|  | Int | -58.404 | (-69.9-46.9) | <0.001 |
|  | Post | 0.086 | (0.06-0.1) | <0.001 |
|  | Gender (female) | -4.286 | (-5.29-3.27) | <0.001 |
|  | Age | -0.12 | (-0.15-0.08) | <0.001 |
| Blood pressure | Pre | -0.002 | (-0.005-0.001) | 0.167 |
|  | Int | 0.522 | (-1.66-2.71) | 0.640 |
|  | Post | -0.006 | (-0.01-0.001) | 0.007 |
|  | Gender (female) | 0.221 | (-1.6-2.04) | 0.812 |
|  | Age | 0.007 | (-0.04-0.06) | 0.808 |
| Cholesterol | Pre | -0.03 | (-0.04-0.01) | <0.001 |
|  | Int | 6.047 | (-0.78-12.87) | 0.083 |
|  | Post | 0.046 | (0.03-0.06) | <0.001 |
|  | Gender (female) | 9.214 | (7.43-10.98) | <0.001 |
|  | Age | 0.121 | (0.06-0.17) | <0.001 |
| VLDL | Pre | 0.007 | (0.003-0.01) | <0.001 |
|  | Int | 1.784 | (0.14-3.42) | 0.033 |
|  | Post | -0.014 | (-0.01;-0.01) | <0.001 |
|  | Gender (female) | -0.108 | (-0.75-0.54) | 0.744 |
|  | Age | 0.016 | (-0.003-0.03) | 0.105 |
| LDL | Pre | -0.017 | (-0.02-0.006) | 0.002 |
|  | Int | -1.776 | (-7.95-4.39) | 0.573 |
|  | Post | 0.033 | (0.01-0.04) | <0.001 |
|  | Gender (female) | 6.900 | (5.58-8.21) | <0.001 |
|  | Age | -0.039 | (-0.07-0.0001) | 0.049 |
| HDL | Pre | 0.022 | (0.01-0.02) | <0.001 |
|  | Int | -8.427 | (-10.16-6.69) | <0.001 |
|  | Post | -0.005 | (-0.008-0.0006) | 0.023 |
|  | Gender (female) | 1.125 | (0.14-2.1) | 0.024 |
|  | Age | 0.092 | (0.06-0.12) | <0.001 |
| Triglycerides | Pre | 0.051 | (0.04-0.06) | <0.001 |
|  | Int | -6.802 | (-11.98-1.61) | 0.01 |
|  | Post | -0.111 | (-0.12-0.09) | <0.001 |
|  | Gender (female) | 6.805 | (5.27-8.33) | <0.001 |
|  | Age | 0.18 | (0.13-0.22) | <0.001 |
| Insulin | Pre | 0.014 | (0.01-0.01) | <0.001 |
|  | Int | -7.207 | (-9.05-5.35) | <0.001 |
|  | Post | -0.017 | (-0.02-0.01) | <0.001 |
|  | Gender (female) | 0.207 | (-0.3-0.71) | 0.427 |
|  | Age | -0.001 | (-0.01-0.01) | 0.885 |
| Glucose-linked hemoglobin | Pre | 0.013 | (0.003-0.02) | 0.007 |
|  | Int | -3.227 | (-8.41-1.96) | 0.223 |
|  | Post | -0.051 | (-0.06-0.03) | <0.001 |
|  | Gender (female) | 4.468 | (2.96-5.97) | <0.001 |
|  | Age | -0.011 | (-0.05-0.03) | 0.643 |
| Fasting glucose | Pre | 0.019 | (0.006-0.03) | 0.003 |
|  | Int | 8.873 | (1.76-15.98) | 0.014 |
|  | Post | -0.095 | (-0.11-0.07) | <0.001 |
|  | Gender (female) | 6.721 | (5.37-8.06) | <0.001 |
|  | Age | 0.119 | (0.07-0.15) | <0.001 |
| Health Care Costs | Characteristics | dy/dx | CI95% | p |
| Direct cost | Pre | 1.153 | (1.12-1.17) | <0.001 |
|  | Int | 1189.518 | (1179.29-1199.74) | <0.001 |
|  | Post | -4.323 | (-4.35-4.29) | <0.001 |
|  | Gender (female) | -100.838 | (-103.32-98.34) | <0.001 |
|  | Age | 1.303 | (1.22-1.38) | <0.001 |

**Obs.:** pre = pre-intervention; int = intervention; post = post-intervention.

## Discussion

Moderate to severe obesity impose important socioeconomic and health burden on individuals, health systems and societies worldwide; therefore, its prevention and treatment may represent substantial impacts on health status, quality of life and utilization of health care resources. Results presented in the study showed reduction in direct health care costs and improvements in multiple health outcomes of cohort of patients who had bariatric surgery in a reference hospital within Brazilian health system, comparing 180-day period pre- and post-intervention through interrupted time series analysis with GEE, including controls for individual characteristics.

In addition, micro-costing technique allowed to identify main health care cost drivers pre- and post-surgery in high complexity institution from the public sector in Brazil. Although there was an initial rise in overall health care costs during the period of bariatric surgery, post-intervention trends were significantly negative.

Pre-intervention results allowed the analysis of cumulative health care costs related to procedures related to comorbidities and monitoring during waiting list for bariatric surgery. Data from health outcomes showed worsening of patients’ health status regarding main comorbidities during the period before the intervention, especially weight gain, hypertension, type 2 diabetes, and dyslipidemia.^23,64^

Lopes et al., in a systematic review and meta-analysis has found that there is important decline in drug prescription and health expenditure related to comorbidity resolution of several obesity-related diseases like arterial hypertension and T2DM. The prevalence of arterial hypertension improved 78% and T2DM improved 92%. In the same study, the authors found that anti-hyperglycemic medication have been discontinued in 93% of diabetic patients after surgery and anti-hypertensive drugs were able to be discontinued in 35% of patients and reduced in 48%.^64^ Similarly, Sussenbach et al., have found resolution of comorbidities like T2DM, arterial hypertension and dyslipidemia higher than 95% at 36 months after surgery, with important reduction in the medical costs in the course of the post-operative period, in relation to expenses for medications, professional care, and examinations in the preoperative period.^41^

Considering results obtained in post-surgery health outcomes, there was improvement of patients’ health status, especially referring to parameters related to diabetes and hypertension. Evidence from previous studies indicate that bariatric surgery for treatment of moderate to severe usually improves general health conditions and quality of life of patients, including control and reduction of effects from comorbidities like hypertension and diabetes, at relatively low cost to the public health system.^15,65^

The mean cost for each unit of decreased obesity health-related outcome was quite reasonable in terms of public health system. The cost of US$ 61.68 per kg of weight loss and above US$ 100 to T2DM biomarkers like glucose-linked haemoglobin and fasting glucose to one unit of decrease, the bariatric surgery shows costs relatively above the drug therapy. In our country, the annual expenditure per patient with treatment for T2DM with medicines reach US$ 259.36 and in the rest of the world range from US$ 1.937 to US$ 13.243 or US$ 63.722 in 35-year lifetime.^6,33,66–68^

Considering cardiovascular risk, with is high related to very expensive costs to health systems, our study has found costs from US$ 102.64 to US$ 982.41 (for triglycerides and Vldl, respectively) per one unit of fall in the blood exam. In our country, the average annual cost in medicines for dyslipidemia is US$ 1.417 to US$ 2.300 and the nutritional and laboratory follow-up is nearly US$ 848.^41^ The data found in our study can be also considered at very low cost, mainly take in place the long-term benefit of reduction on the risk of mortality, morbidity, and high cost.^69,70^ Cholesterol was the only biochemical marker showing different pattern from expected; however, it was also not statistically significant. Otherwise, the results confirm trends in enhancement of patients’ health status and reduction in overall health care costs post-bariatric surgery, after controlling for patients’ characteristics.^15^

To date, traditional observational studies perform assessment of costs involved in procedures and/or measurement of effectiveness of bariatric surgery using mean pre- and post-intervention data for comparison of few health outcomes. However, it comprises an approach that neglects the identification of trends in patients’ costs and/or health outcomes during pre- and post-surgery at individual level.

The present study provides information with additional precision on impacts of bariatric surgery considering direct costs and multiple health outcomes related to comorbidities of moderate to severe obesity. The ITS and GEE modelling approaches allowed to assess marginal effects of bariatric surgery in evolution of costs and health outcomes, including correction for individual-level characteristics which reduces potential bias of estimation.

Therefore, our results suggest potential causal relationship between bariatric surgery and improvement of health outcomes in patients with severe to moderate obesity. Considering that patients were followed during similar periods before (waiting list) and after (follow-up) bariatric surgery for comparison of information pre- and post-intervention, we obtained robust results indicating that bariatric surgery was efficient intervention to interrupt worsening of patients’ health conditions, and to promote improvements in health outcomes related to hypertension and diabetes in association with reduction of direct health care costs after treatment, findings in accordance with previous evidence.^15,19–21,23,24^

Due to lack of previous evidence regarding impacts of bariatric surgery based on pre- and post-intervention costs and multiple health outcomes using micro-costing approach at individual-level, the robust results provided by our estimates may provide real-world foundations for public policy decision making within Brazilian national health system, especially referring to intervention with significant potential for reduction of burden of diseases related to obesity.^71,72^

The analysis was conducted on small sample of patients with moderate to severe obesity within one Brazilian hospital of reference who were on the waiting list for bariatric surgery, followed from 6-month period before and after the bariatric surgery, extracted from a cohort of approximately 2,000 patients followed during 12 or more years. The selection of sample for analysis using the interrupted time series analysis with GEE was based on eligibility of patients, considering the existence of complete information registered in electronic medical records encompassing periods pre- and post-intervention and the need for micro-costing approach using data obtained manually for each patient. However, in view of promising results obtained through application of the statistical strategy for assessment of trends regarding direct health care costs and health outcomes, additional information may be obtained for further investigation of the subject.

## Conclusion

Bariatric surgery represents an effective intervention for treatment of moderate to severe obesity with extensive benefits regarding health promotion and reduction of burden of disease. Trends in direct costs and multiple health outcomes showed post-intervention improvements in patients’ health status and reduction of health care needs of individuals.

## Data Availability

The data that support the findings of this study are available from the corresponding author JAOT, upon reasonable request. Restrictions apply to the availability of patients medical datasheet, which were used under hospital permission for this study.

https://www.dropbox.com/home/Doutorado/Paper%202%20-%20CE%20Cir%20bar%206m%20long

## Acknowledgements

The study was financed in part by the Coordenação de Aperfeiçoamento de Pessoal de Nível Superior - Brasil (CAPES) - Finance Code001 - 88887.368403/2019-00.

The authors also thank the contributions from the Department of Gastroenterology, the Department of Obstetrics and Gynecology, the Hospital of Clinics (HC-FMUSP), Mr. Vilson Cobello Júnior (Coordinator of Technology of Information at HC-FMUSP), and Mr. Jacson Venâncio de Barros (Director of Informatics Department of the Brazilian Public Health System, DATASUS).

## Data availability statement

The data that support the findings of this study are available from the corresponding author JAOT, upon reasonable request. Restrictions apply to the availability of patients’ medical datasheet, which were used under hospital permission for this study.

